# Is there a link between scoliosis and headache disorders-related symptoms?

**DOI:** 10.1101/2023.10.22.23294683

**Authors:** Miriam Faes, Claudia Milella

**Affiliations:** Department of Human Neuroscience, University “La Sapienza”, Roma

## Abstract

**Background:** Scoliosis is a complex deformity of the spine in all three planes. Its multifactorial etiology is not fully clear: it results from genetic, endocrine and muscular factors connected with biomechanical causes. The most common form is adolescent idiopathic scoliosis (AIS), usually detected during the pubertal growth spurt. It affects 2–3% of adolescents with some restrictions to their daily life, being a risk factor for different impairments to health-related quality of life (HRQoL). With advances in this field of study, more attention has been given to the perception of deformity and the correlated symptoms of some comorbidities.

Among different aspects, several studies are focusing on the potential of scoliosis to cause postural control impairments.

To date, in literature, a plausible relationship between scoliosis and headache disorders is sparsely studied not only in adult patients but also in adolescents. The diagnosis of the different types of headaches remains a challenge even because they are similar, having several symptoms that can be considered the same.

**Objectives:** The aim of this scoping review is to provide an analysis of the current state of the art on scoliosis and the possible correlation with headache-related symptoms.

**Methods:** Research was conducted through all publications written in English about this topic in accordance with the PRISMA extensions for Scoping Reviews (PRISMA-ScR). Studies were included if they had met the following inclusion criteria: population affected by primary scoliosis, no limits of age and gender. No study design, publication type and data restrictions were applied. Medline (PubMed), gray literature databases were searched up to May 2023. Two reviewers managed to screen independently all titles, abstracts and free full-text studies for inclusion. A data collection form was developed in order to extract the characteristics of the studies included.

**Results:** In recent years, greater attention has been given to the perception of the deformity and to the correlated symptoms of different comorbidities. A few studies focus on the potential of scoliosis for causing postural control impairments that can pave the way to headache. A plausible relationship between scoliosis and headache disorders is sparsely studied not only in adult patients but also in adolescents. Data on percentage of headache as a comorbidity are present in 7 studies, which met the elegibility criteria for the review. Heterogeneity of outcome measures and participants in the studies caused the impossibility to pool together the results in a statistical form. Therefore, it was decided to report them in narrative form.

**Conclusion:** The data included seem not sufficient to guarantee a reasonable quality of analysis. Specifically, there is a lack of systematic research with a focus on the presence of a possible correlation between scoliosis and headache. The significant heterogeneity of symptomatic dimensions and sometimes their overlap occur in the cephalalgic patient and it complicates the interpretation of the data. It seems necessary to continue research in order to investigate through a questionnaire -Midas adapted-that can be submitted to scoliotic patients and offer a good methodological perspective. Moreover, in case of the presence of headache, a detailed analysis could also be significant through their symptom diary.

## INTRODUCTION

### Background

Scoliosis is a three-dimensional deformity of the spine that shows an abnormal lateral curvature, greater than 10° Cobb, and the rotation of the vertebrae within the curve (1). Physical examination of the back, an X-ray, spinal radiograph, CT scan or MRI are helpful to diagnose scoliosis and the deformity is measured by the Cobb Method. Scoliosis can make the hips, shoulders and ribs appear uneven and there can be a curve typically called a C curve or multiple curves, typically called a S curve. When vertebrae twist, one shoulder can blade protrude (Scoliosis Research Society). The causes of scoliosis vary and its multifactorial etiology (2) can be used to classify each case as congenital, neuromuscular, syndrome-related, spinal curvature due to secondary reasons or idiopathic scoliosis. The latter is the most common form, it occurs in the 80% of the patients of all ages and can progress during a rapid period of growth (1).

Adolescent Idiopathic Scoliosis (AIS) is diagnosed between the ages of 10 and 18 and affects 4% of adolescents worldwide (Scoliosis Research Society). It is suspected to be present in 2 to 4 % of children and adolescents (3). In current literature the prevalence differs across the countries from 0.47 % to 12% and the leading estimate between 2% and 3% is widely accepted (4).

The ratio of female to male AIS diagnoses is ranging from 1.5:1 to 3:1(5). Females show a more rapid curve progression and are eight times more likely to be treated surgically (6).

The severity of AIS is described by the number of degrees and cases range from mild (10 to 20°Cobb), to moderate (21 to 35°), to severe (41° and above). Treatments are on a case-by-case basis due to age, curve magnitude and risk of progression; they include observation, specific types of physical therapy, orthotic management with various braces and surgical correction (Scoliosis Research Society).

In adults degenerative scoliosis is a consequence of altered biomechanics in a spine with previous idiopathic scoliosis (treated when adolescents with surgery or with conservative treatment) or an asymmetric degeneration of the disc and facet joint and a *de novo* deformity.

Although AIS can progress during growth and the surface deformity can worsen, it does not usually result in pain or neurologic symptoms (7). Among the complications, the ones that are widely recognised are breathing problems, especially in severe scoliosis, and back pain (8). Low back problems can be present as chronic in association with neck pain in adult women in particular (9) and in patients with untreated abnormal curves (10) or during curve progression. In adolescents low back pain is not uncommon (11).

The significant restrictions in everyday activities are under attention thanks to advances in the study of health-related quality of life, as deformity can be regarded as a risk factor for this type of impairment (12).

Besides, many people with scoliosis develop pain in other parts of their bodies: they generally show reduced or delayed muscle mass which is a mismatch between skeletal growth and muscular-ligamental maturation. Among other interesting perspectives, various studies have focused on co-existent abnormalities in adolescents but knowledge about the neurodynamic functions of the nervous system is still limited (13).

As far as headache disorders are concerned, in the IHS, International Headache Society-3rd edition (2018) no links can be traced between scoliosis and headache. No data are present in the literature but these common debilitating disorders can be reported with a wide range of potential accompanying symptoms by patients with AIS.

### Objectives

This scoping review will attempt to highlight an overview of the current concept of AIS and to find some evidence to guide future research in discussing the relationship between this spinal deformity and the different types of headache disorders.

## METHODS

The results of this scoping review were reported using the Preferred Reporting Items for Systematic Reviews and Meta-Analyses (PRISMA 2020) extension for Scoping Reviews (PRISMA-ScR) Checklist (14).

### Eligibility Criteria

The research was conducted through various publications written in English. Studies were selected from any year of publication with the following inclusion criteria: population affected by primary scoliosis, no limits of age and gender, and any type of headache disorders.

### Information sources

The research was conducted on the PubMed database (Medline), started in January 2023, and ended in May, 2023.

### Search

The research strategy was designed according to the specific settings of the reference database (PubMed. It was developed using a PEO model of the research query: free terms or synonyms were used, in combination with the Boolean operators (AND, OR); no filters were applied by year of publication.

Furthermore, a manual search was conducted through the different bibliographies of the articles reviewed in order to implement additional studies eligible after the initial screening.

The following research strategy was used:

P(*patients*): people affected by headache disorders

E (*exposure*): scoliosis

O (*outcome*): /

(scoliosis OR “spinal deformities” OR “spinal abnormalities” or “idiopathic scoliosis”) AND (headache OR migraine OR cephalalgia OR “tensive headache”)

((“scoliosis”[MeSH Terms] OR “scoliosis”[All Fields] OR “scolioses”[All Fields] OR “spinal deformities”[All Fields] OR “spinal abnormalities”[All Fields] OR “idiopathic scoliosis”[All Fields]) AND (“headache”[MeSH Terms] OR “headache”[All Fields] OR “headaches”[All Fields] OR “headache s”[All Fields] OR (“migrain”[All Fields] OR “migraine disorders”[MeSH Terms] OR (“migraine”[All Fields] AND “disorders”[All Fields]) OR “migraine disorders”[All Fields] OR “migraine”[All Fields] OR “migraines”[All Fields] OR “migraine s”[All Fields] OR “migraineous”[All Fields] OR “migrainers”[All Fields] OR “migrainous”[All Fields]) OR (“headache”[MeSH Terms] OR “headache”[All Fields] OR “cephalalgia”[All Fields] OR “cephalalgias”[All Fields]) OR “tensive headache”[All Fields]))

### Selection of sources of evidence

The process for selecting sources of evidence was performed individually by a single reviewer (MF), under the supervision of a second author (CM). The *full texts* of the identified studies were searched for further evaluation according to the pre-set eligibility criteria.

### Data charting process

Data were collected by a single reviewer (MF) through *full text* reading of included studies, in some cases added and always supervised by a second author (CM).

### Data items

For each article the following were extracted:

- *First author’s surname and year of publication;*
- *article’s title and publication;*
- *objective of the study;*
- *population (sample size, demographic characteristics of the sample);*
- *diagnostic method of scoliosis*
- *results of the study*
- *conclusions of the study*.

### Critical appraisal of individual sources of evidence

The *JBI Critical Appraisal Checklist for Analytical Cross-Sectional Studies*, developed by the Joanna Briggs Institute (2017), was used to assess the risk of bias in each of the studies included in the review. This checklist is made up of six items:

1. *Were the criteria for inclusion in the sample clearly defined*?
2. *Were the study subjects and the setting described in detail?*
3. *Were objective, standard criteria used for measurement of the scoliosis?*
4. *Were links with headaches identified?*
5. *Were the outcomes measured validly and reliably?*
6. *Was appropriate statistical analysis used?*

For each question, the possibilities for answering are as follows: “*Yes*”, “*No*”, “*Unclear*” and “*Not applicable*”.

The evaluation was performed by the two authors.

### Synthesis of results

It was decided to report them in narrative form.

This manuscript has been seen and approved by all listed authors.

The authors have declared no competing interests.

### Data Availability

All data produced in the present study are available upon reasonable request to the authors.

## Results

PRISMA Scr

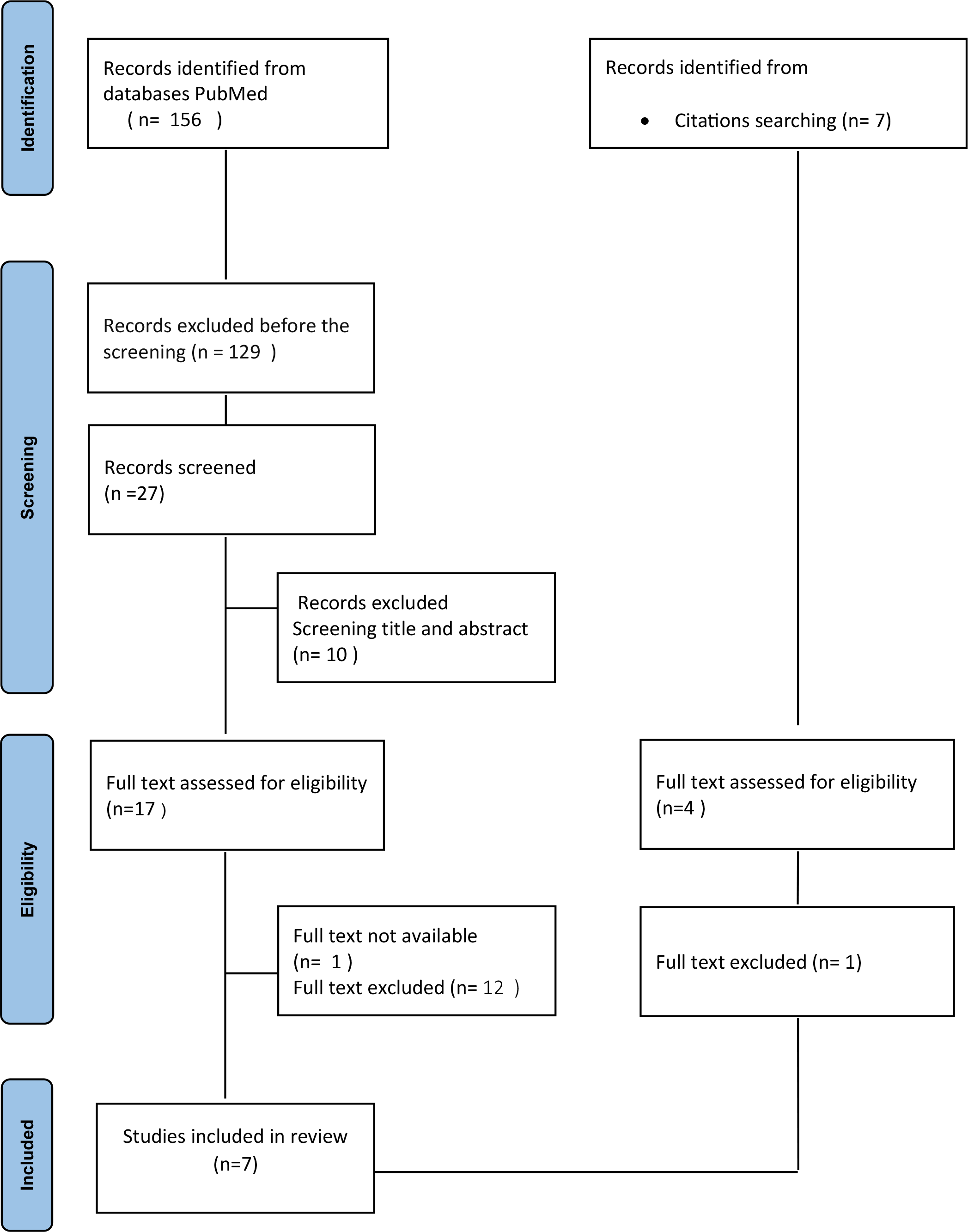

The articles selected for this review are the following:

1. Brox J.I., Lange J.E. (2014)
2. Guyot M.A., Agnani O., Peyrodie L., Samantha D., Donze C., Catanzariti J.F. (2016)
3. Yamamoto T., Narai S. (2004)
4. Uneri A., Polat S., Aydingoz O., Bursali A. (2009)
5. Misterska E., Głowacki J., Okręt A., Laurentowska M., Głowacki M. (2017)
6. Freidel K., Petermann F., Reichel D., Steiner A., Warschburger P., Weiss HR. (2002)
7. Du C., Yu J., Zhang J., Jiang J., Lai H., Liu W., Liu Y., Li H., Wang P. (2016)

Scoliosis is a complex deformity of the spine in all three planes with a multifactorial etiology that presents its most common form as adolescent idiopathic scoliosis (AIS) (15). It affects 2–3% of adolescents with some restrictions to their daily activites and represents a risk factor for impairments in health-related quality of life (HRQoL).

In recent years, greater attention has been given to the perception of the deformity and to the correlated symptoms of different comorbidities. A few studies focus on the potential of scoliosis for causing postural control impairments that can pave the way to headache. A plausible relationship between scoliosis and headache disorders is sparsely studied not only in adult patients but also in adolescents. Data on percentage of headache as a comorbidity are present in 7 studies.

In the majority of cases scoliosis is asymptomatic. The scientific literature on pain offers various case studies on patients whose curve varies in severity and is treated with different types of braces.

Each patient has to face this pathology that undermines the column, i.e. the supporting structure of the body. In particular, Scoliosis Research Society confirms that pain is not uncommon and urges a better evaluation of each patient’s emotional response, especially in the delicate moment of growth and pubertal development. The impact of spinal deformities on health-related quality of life has been considered with greater attention since the 1980s, given that they can lead to relevant consequences (16). For scoliosis < 20° Cobb, limitations to daily activities have never been described (17) but as the disease worsens, the repercussions on the quality of life may become increasingly more impactful. In his study, Du reports that pain in the head and neck is complained by 15 % of young patients.

It should not be overlooked how much the evolution of the curve is linked to strictly individual factors which, over time, could give rise to or could be accompanied by, sensory and emotional experiences, perceived in a personal and more disabling way than others.

The study by Misterska takes into consideration musculoskeletal deformities that could influence the perception of pain in the low back and in the neck, two body districts where symptoms of pain can be evaluated more widely (9).

As regards adult patients, there are studies that provide a long-term follow up. Specifically, in 2014, Brox assessed quality of life related to comorbidities and health conditions in patients in their fourth decade of age, with scoliosis previously treated with braces (18). The sample consists of 496 patients who had the Boston brace; at follow-up after 23 years, 390 people partecipated: 361 women and 29 men, with an average age of 39.2 years (± 4.6 years). During the investigations, 122 patients reported, at least, one comorbidity that influenced their perception of life among asthma, **migraine**, lower limb problems, depression, anxiety, lumbar hernia, neck pain and widespread pain. The data on migraine that emerge from this cross-sectional study appear significant, despite the diagnostic difficulty of migraine. Actually, 13 patients reported migraine in a specific way. New tool, different from EuroQol, used by Brox, should discriminate the comorbidities in order not to underestimate their effect.

Restricting data on female population, Freidel and colleagues examined 226 scoliotic patients divided into two groups by age: from 17 to 21 and over 21 years of age. To the first group, the younger one, the Berner Questionnaire for well-being (BFW) was administered (19). This questionnaire can detail 6 scales about positive attitude towards life, awareness of problems, self-esteem, tendency to depression and sense of satisfaction in life, “body complaints and reactions” as an item including diversified symptoms, such us stomachache, palpitations, loss of appetite, sleep disturbances, feeling of heat and **headache**. Compared to the group of age-matched control, girls between 17 and 21, showed their negative preception of life and the score in this specific item, emerged as higher, with a 2.29 compared to 1.89 (SD).

From the analysis of the responses to the Short Form Health Status Survey (SF 36) in the study by Freidel, the second group of 80 people over the age of 17 highlighted the score of average bodily pain slightly lower than the control sample.

In this study stomachache and headache are considered underestimated and they can be considered non-symptoms; although they are present in young patients with scoliosis, the data do not support the hypothesis that this dysmorphism may represent a possible risk factor and concomitant with pain and/or headache; the same patients with a worsening scoliosis and increasing age reported they had faced significant physical problems in their life.

Although the study helps to spread the idea about the importance of a more attentive approach to psychosocial difficulties and the support to an individual perspective towards pain in the patient with AIS the conclusions of the article cannot offer further details because the classification of headache appears to be rather generic.

After having verified the research question also in gray literature, the study by Yamamoto and Narai proved to be interesting. It explains the higher incidence of scoliosis in children with tension-type headache symptoms when compared to the general population (20). The analysis was set in Japan on a sample of 71 children with headache. Out of these patients, 5 would be evaluated and diagnosed later for scoliosis. Within the 11 cases of scoliosis in the study, 3 girls reported also headache. The authors’ suggestion to pediatricians seems valid after many years in order to pay attention to the alignment of the spine in children with headache: the anomalies in the posture of the trunk can, in fact, be factors for triggering headache.

Regarding posture, the recent study by Larni explains the anomalies in biomechanics and postural control variables in adolescents with scoliosis, their postural instability and the difficulty in the integration mechanisms of data (21). The patients have deficits in muscular effort and counterbalance anti-gravity forces in the attempt to maintain a better and more correct position. These details could provide a plausible correlation with tension headache. This type of headache can also depend on continuous involuntary contractions of the neck and shoulder muscles. More common in female patients, it mainly affects individuals who maintain a prolonged incorrect position.

Scoliosis can be located in different areas of the spine; in some patients it is possible also to note a position of the head that is not symmetrical with respect to the pelvis. In literature there are some studies on the greater frequency of altered posture of the head and in particular on the forward head posture (FHP) in patients with scoliosis (22).

It is also worth considering the fact that the severity of the curve and various treatment attempts could exacerbate the painful sensations and symptoms related to headache. On the other hand, investigations in the field have concluded that neck pain is simply a symptom of a migraine attack. In 2006 De Las Peñas hypothesized that dysfunctions in the cervical muscles and the presence of active and latent points can trigger migraine attacks (23) and also contribute, if not correctly managed, to the transformation of migraine from episodic to chronic (24).

Various studies have drawn their attention on the incidence of cervical disorders in scoliotic patients surgically treated. If its presence were confirmed in larger numbers of scoliotic patients, reference could be made to the probable relationship between cervical disorders and cervicogenic headache.

Many researchers have stated that scoliosis patients suffer from neck pain (NP) and low back pain (LBP), more frequently when compared to the healthy population (25) and have focused on groups of individuals treated conservatively, both with and without brace. In the comparative analysis by Misterska and colleagues (2017), the majority of women with scoliosis, that is the 83.84%, treated with the Milwaukee brace and monitored in a follow up after 23 years, reported a perception of disability from mild to serious. In the control group of healthy women, only 64.29% reported a limited functionality. The researchers used the questionnaire to deduce the results thanks to Neck Disability Index (NDI), that investigates 10 sections of everyday life with questions about intensity of pain, quality of sleep, personal care, concentration, ability to work, drive, read and play and a peculiar item dedicated to headache.

For the question of this review, it is considered appropriate to select only the outcomes obtained at the NDI. In fact, the reported mean of 11.66 ± SD 7.43 in the group of women with scoliosis compared to the mean of 4.38 ± SD 4.02 of the control group may offer further inspiration. Although the score is never separated and cannot be studied individually, it suggests some consistent responses with the research question, albeit limited. Misterska concludes that people with scoliosis and treated with braces in adolescence, show significant limitations in daily activities and higher levels of pain and disability related to NP and LBP when compared to the control group.

Until 2016, the cervical spine had not been studied extensively in patients with AIS: it was Guyot to orient his research to altered proprioception in adolescents with scoliosis (26). In his study the 30 AIS patients between 15.5 ± 1.5 years, with 24.8 ± 9.5 °Cobb, were evaluated by the Cervicocephalic relocation test, after active rotation without visual control, performing 10 rotations of the cervical spine, first to the right and then to the left. Comparing their results with those of the control sample (14 adolescents without scoliosis aged 14.6 ± 2.0 years), they demonstrated a deficient cervical proprioception. Moreover, adolescents with AIS showed a loss of cervical stabilization during the rotation of the head and a greater reliance on visual afferents in order to orient their cervical spine. This anomaly is correlated with a worse prognosis of AIS and is implicated in the onset of headache and balance disorders.

As regards other types of headache that may be present in patients with scoliosis and be correlated with the same deformity, the study by Uneri brings into focus vestibular migraine (27). Vestibular disorders and migraines are characterized by a high prevalence in general population; there are some studies and in particular Cortes-Perez (28) which highlight the association between scoliosis and alterated vestibular morphology (29).

In the study by Uneri, his sample consisted of 7 members of 3 families with scoliosis of varying severity and vestibular dysfunctions accompanied by migraine. The description of each case is thoroughly presented but the number of patients is small and the evolution in the etiopathogenesis of headache do not offer further perspectives on the topic. The association between dizziness and headache appears insufficient to possibly diagnose migraine vestibulopathy in subjects suffering from scoliosis.

In conclusion, the evaluation of the vestibular system could prove significant for a comprehensive analysis of musculoskeletal deformity and its impact on patients’ life. Vestibular treatment could further improve posture and reduce the risk of curve progression. The lack of data and the limited percentage of patients with scoliosis and vestibular problems complicate the analysis.

## Conclusion

In this scoping review we attempted to answer the question mapping first of all the most recent scientific literature on the key concepts of scoliosis and headache in order to broaden its knowledge for professional purposes. Through the years little research has been done on the relationship between scoliosis and headache and especially in the Italian context. The reason could be related to the heterogeneity of symptoms and the attention that is paid to evolution and progression of the curve rather than to symptoms associated with it. The data from the 7 studies included seem not sufficient to guarantee a reasonable quality of analysis. Specifically, there is a lack of systematic research with a focus on the presence of a possible correlation between scoliosis and headache. The significant heterogeneity of symptomatic dimensions and sometimes their overlap occur in the cephalalgic patient and it complicates the interpretation of the data.

Furthermore, the groups of eligible patients received different types of treatment for scoliosis and this aspect does not allow a valid analysis of the results. A further limitation can be found in the large difference in type and length of the treatment and its follow-up, making it difficult to offer an objective comparison. Despite limitations, the review clarified in part some aspects relating plausible relationships between scoliosis and tension or cervicogenic headache. The increase in cases of the musculoskeletal deformity and the attention to the well-being of patients can suggest the necessity of further studies. Moreover, it would be necessary to systematically study the impact of scoliosis in women and the quality of their lives, evaluating headache as contingent element or as possible risk factor.

Factors such as stress and alterations in psychological well-being should be investigated aiming to prove triggers to postural and proprioceptive deficits in the different stages of the curve and the onset of any headache disorders.

In conclusion, it seems necessary to continue research in order to investigate through a questionnaire -Midas adapted-that can be submitted to scoliotic patients and offer a good methodological perspective. In fact, in the case of the presence of headache, a detailed analysis could also be significant through their symptom diary.

